# Dynamic Risk Trajectories for Sudden Cardiac Arrest: The Role of Recurrent Cardiovascular Events

**DOI:** 10.1101/2025.11.13.25340202

**Authors:** Marita Knudsen Pope, Harpriya Chugh, Thien Tan Tri Tai Truyen, Marco Mathias, Audrey Uy-Evanado, Honghuang Lin, Dan Atar, Nichole Bosson, Kyndaron Reinier, Emelia J. Benjamin, Sumeet S. Chugh

**Affiliations:** Center for Cardiac Arrest Prevention, Smidt Heart Institute, Cedars-Sinai Medical Center, Los Angeles, California, USA; Department of Cardiology, Oslo University Hospital, Ullevaal, and the Institute of Clinical Medicine, University of Oslo, Oslo, Norway; Department of Medicine, University of Massachusetts Chan Medical School, Worcester; Los Angeles County Emergency Medical Services Agency; Boston University and National Heart, Lung, and Blood Institute’s Framingham Heart Study, MA; Section of Cardiovascular Medicine, Department of Medicine, Boston Medical Center, Boston University Chobanian & Avedisian School of Medicine, MA; Department of Epidemiology, Boston University School of Public Health, MA

## Abstract

**Background:** Emerging evidence suggests that dynamic risk assessment may enhance sudden cardiac arrest (SCA) risk stratification. While cardiovascular events, including acute coronary syndrome (ACS) and heart failure (HF) hospitalization, are associated with increased SCA risk, the impact of recurrent events on subsequent SCA risk in a contemporary, real-world population is unknown. This study aimed to assess whether patients with a first-time ACS or HF hospitalization who experience a recurrent cardiovascular event have higher risk of SCA compared to those who do not.

**Methods:** The Observational Study of Cardiac Arrest Risk (OSCAR) is a prospective cohort study with adjudicated SCA outcomes. In the current study, patients who survived a first ACS or HF hospitalization were categorized into index ACS or HF cohorts. Participants were followed for recurrent cardiovascular events and SCA. Associations between recurrent events and SCA were assessed using Cox models with recurrent event modeled as a time-dependent variable.

Findings were validated in the Framingham Heart Study (FHS).

**Results:** In the OSCAR discovery cohort, 2946 patients experienced an index ACS event. The incidence rate of SCA was higher following a recurrent ACS event than without (3.70 vs 1.28 per 100 patient-years). Recurrent ACS event was associated with a significantly higher risk of SCA (adjusted HR 3.15, 95% CI 2.06-4.83, p<0.0001). A total of 6711 patients experienced an index HF hospitalization, and the incidence rate of SCA was higher following a recurrent HF event than without (1.35 vs 0.97 per 100 patient-years). Recurrent HF hospitalization was associated with a significantly higher risk of SCA (HR 1.81, 95% CI 1.46-2.26, p<0.0001).

In the FHS validation cohorts a recurrent event during follow-up was associated with a significantly higher risk of SCA in the ACS cohort (HR 2.85, 95% CI 1.66-4.90, p=0.0002), but the association was not statistically significant in the HF cohort (HR 1.49, 95% CI 0.73-3.03, p=0.27).

**Conclusion:** Recurrent ACS event was associated with more than threefold higher risk of SCA, and a recurrent HF hospitalization with 80% higher risk of SCA. These findings suggest that dynamic clinical trajectories of recurrent cardiovascular events may inform management and prevention of SCA.

**Clinical Perspective:** *What is new?:*

- In patients with a first acute coronary syndrome (ACS), those with a recurrent ACS event during follow-up had a threefold higher risk of sudden cardiac arrest (SCA) compared with patients without recurrence.
- Among patients hospitalized for heart failure (HF) for the first time, those with a subsequent HF hospitalization had nearly double the risk of SCA compared with those without recurrence.

*What are the Clinical Implications?:*

- These findings indicate that individuals with recurrent cardiovascular events are at higher risk of sudden cardiac arrest.
- These dynamic risk trajectories may inform management of these patients as well as improved prevention of sudden cardiac arrest.
- Future prospective studies are needed to test these findings in larger and more diverse populations.

## Introduction

Sudden cardiac arrest (SCA) is a major public health challenge, resulting in substantial loss of life each year. In the United States alone, more than 350,000 individuals experience SCA annually, with survival rates remaining below 10%^1^. Current SCA risk prediction primarily relies on static measures, with left ventricular ejection fraction (LVEF) <35% serving as the principal selection criterion for primary preventive implantable cardioverter-defibrillator (ICD) implantation^2,3^. However, this approach has limited specificity and sensitivity; advances in heart failure (HF) management have led to a reduction in SCA rates among patients with reduced LVEF^4^, and the majority (>70%) of patients who experience SCA in the community have LVEF greater than 35%^5^.

Consequently, there is a critical need for improved risk stratification strategies. Emerging evidence suggests that dynamic risk assessment, accounting for changes in vulnerable clinical substrate over time, may improve SCA risk stratification^6,7^.

Acute coronary syndrome (ACS) and heart failure (HF) are well-established pathophysiological conditions that predispose to SCA^8^. Patients who experience recurrent cardiovascular events, including new ACS episodes or HF hospitalizations, are at elevated risk of major adverse cardiac events^9–12^. Recurrent cardiovascular events may reflect progression of underlying myocardial injury, ischemia, electrical instability, or worsening ventricular remodeling, all of which can contribute to increased susceptibility to malignant arrhythmias. However, the association between recurrent cardiovascular events and the risk of SCA remains insufficiently characterized. A few post hoc analyses from randomized controlled trials (RCT) have suggested an increased risk of sudden death among ACS patients who experience recurrent cardiovascular events^13–15^. In patients with HF, some studies have reported a higher risk of malignant arrhythmias in those with recurrent events; however, these studies were restricted to ICD recipients and did not specifically evaluate SCA as an outcome^16,17^.

In the current study, data from the large, prospective Observational Study of Cardiac Arrest Risk (OSCAR) cohort were used to investigate the associated risk of SCA in patients with a first ACS or HF hospitalization who experienced a recurrent cardiovascular event compared with those who did not. The findings were independently validated in the Framingham Heart Study (FHS).

## Methods

### Discovery cohort

#### Study design

For the discovery cohort, we identified patients from the OSCAR database. The study has been described in detail previously.^18^ In brief, the OSCAR cohort is an electronic health records (EHR) study of Los Angeles residents with at least one patient encounter with the Cedars-Sinai Health System in two consecutive years during the period from 2016 to 2020. EHR data were obtained from all outpatient and inpatient encounters across the health care levels from year 2012.

For the current study, we included patients aged ≥35 years with a first hospitalization for ACS or HF after enrollment in OSCAR in two distinct cohorts: an index ACS cohort and an index HF hospitalization cohort, see figure 1a. We only included patients who survived to hospital discharge from their index event to ensure a population truly at risk of experiencing a recurrent event. We excluded patients with a prior history of the same cardiovascular event type from the given cohort. Patients could contribute to both cohorts. We then prospectively followed patients for recurrent event of the same index event type, and for the outcome of SCA. Only the first occurrence of a recurrent event was considered. Time at risk ended at time of SCA, death or at the end of follow-up. There were no criteria for survival time after a recurrent event. At the time of data extraction, follow-up data were available until September 2024, making the maximum follow-up time 7.75 years.

**Figure 1a).**
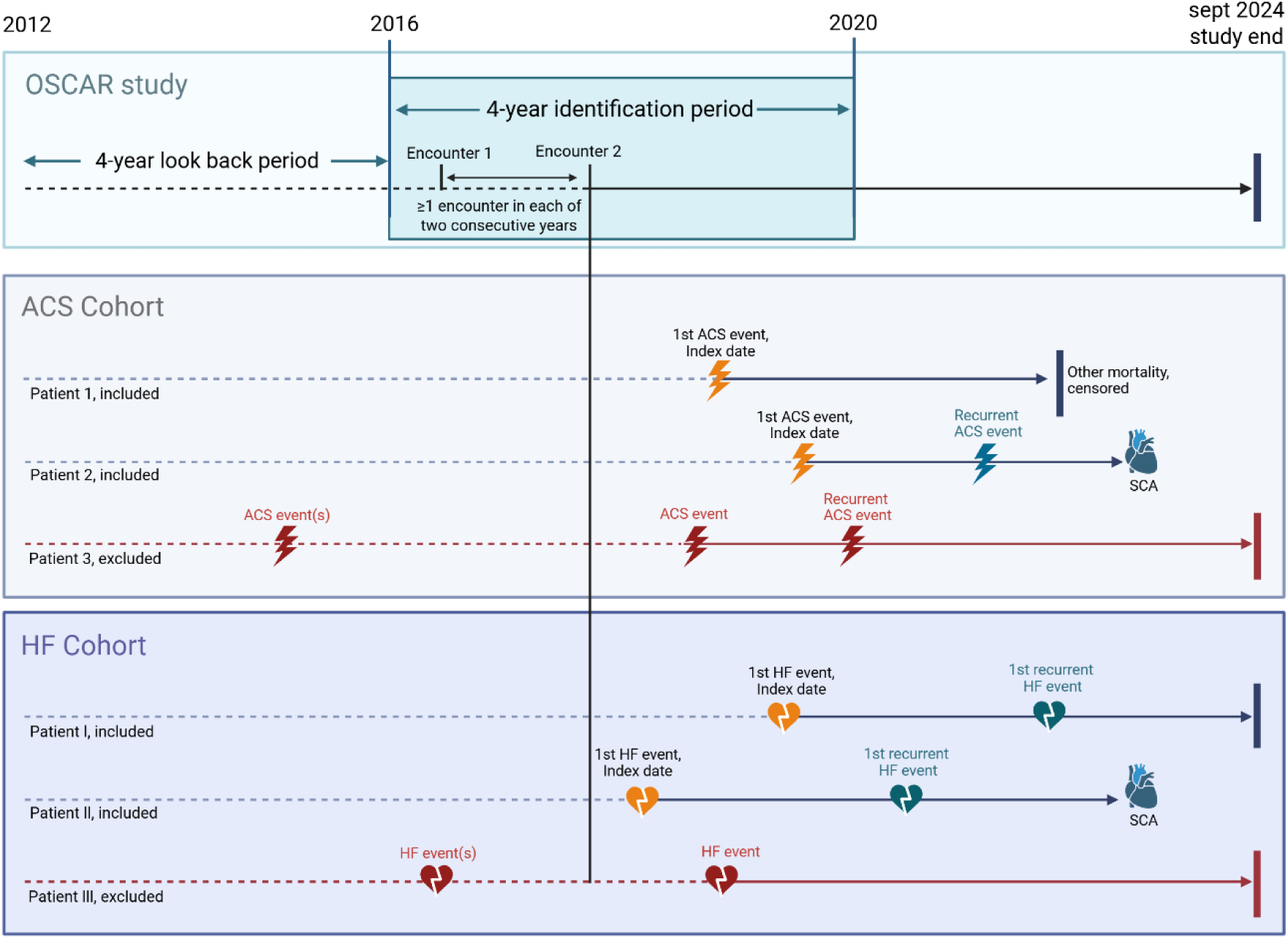
Illustration of the study design. a) Identification of patients from the OSCAR study to include in the ACS and HF hospitalization discovery cohorts. Enrollment in OSCAR required at least one patient encounter within the Cedars-Sinai Health System in two consecutive years. Abbreviations: ACS, acute coronary syndrome; HF, heart failure; OSCAR, Observational Study of Cardiac Arrest Risk; SCA, sudden cardiac arrest

**Figure 1b).**
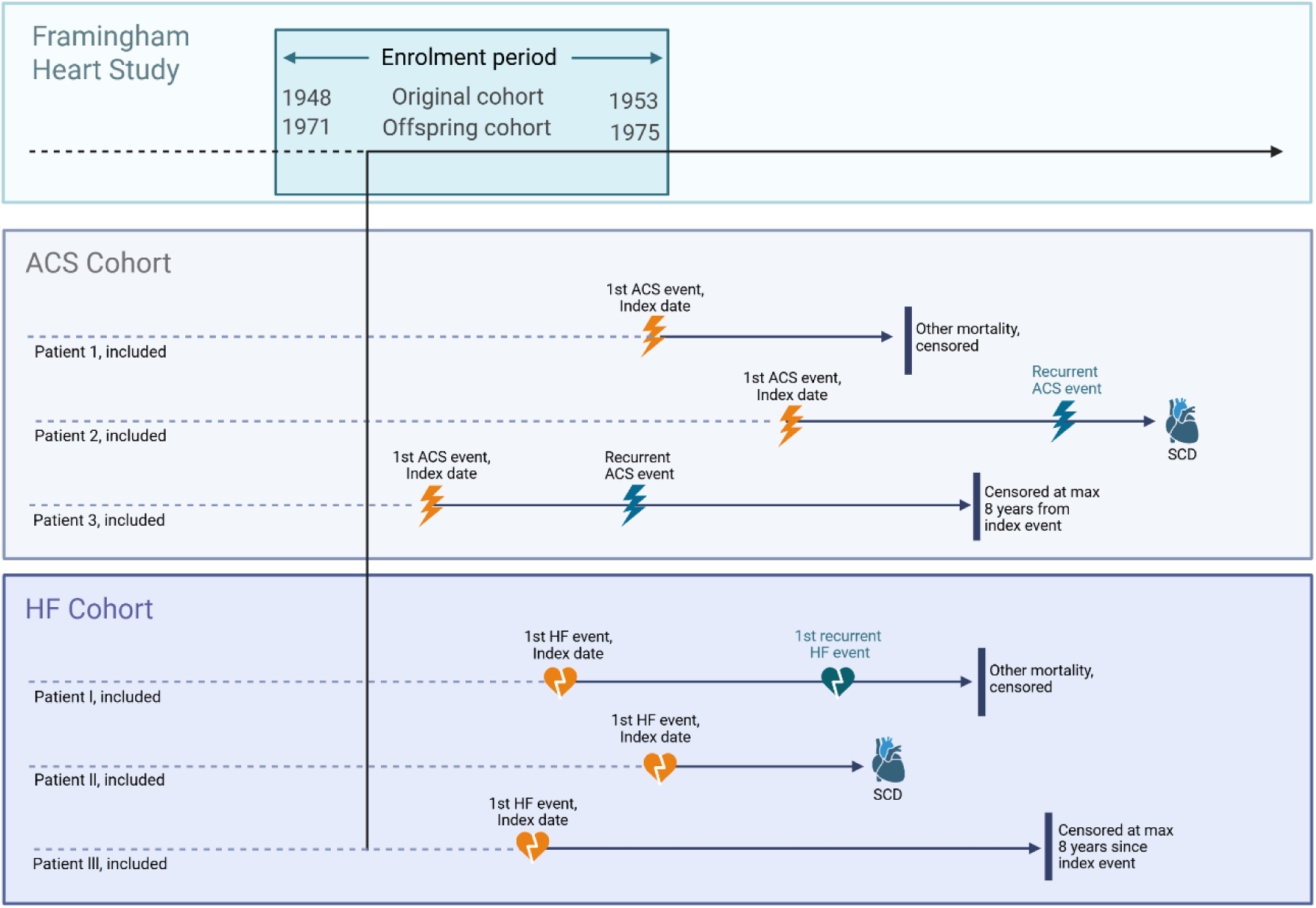
Identification of patients from the FHS study to include in the ACS and HF hospitalization discovery cohorts. Abbreviations: ACS, acute coronary syndrome; FHS, Framingham Heart Study; HF, heart failure; SCD, sudden cardiac death.

#### Variables and clinical event definitions

We retrieved demographics, risk behavior, anthropometrics, and clinical comorbidities from the EHR during the baseline period. Clinical events and treatment during follow-up were defined by appropriate ICD-10 diagnostic and procedure codes as well as CPT codes, and an ACS was defined as a hospitalization for unstable angina (UA) or myocardial infarction (MI), and HF hospitalization a hospitalization where the primary ICD-code was HF (ICD-10 code list in Supplementary Table 1a and b).

We identified cases of presumed SCA from the regional emergency medical services (EMS) database. Thereafter, trained research physicians conducted a careful case adjudication process using a standardized method as well as all available data, including medical records,

EMS reports, and death certificates^18^. SCA was defined as a sudden and unexpected loss of pulse due to a likely cardiac cause,^19^ and patients whose SCA occurred due to trauma, substance use, or natural non-cardiac causes were excluded. Survivors of SCA (n=26, 5.1% of total SCA) were included.

### Validation cohort

#### Study design

For the validation cohort, we extracted data from the Framingham Heart Study (FHS). FHS is a community-based epidemiological study conducted in Framingham, Massachusetts. The study methodology has been described in detail previously^20,21^. The study was initiated in 1948 with an Original Cohort of 5209 participants and was extended with an Offspring Cohort of 5124 participants in 1971. All participants underwent an initial interview and physical examination, followed by follow-up examinations approximately every two to four years.

For the current study, we categorized participants with an index ACS or HF hospitalization during follow-up into two corresponding cohorts, as done in the discovery cohort (see figure 1b). Included participants were ≥35 years old at time of index event. As data on hospital length of stay were not available in FHS, survival to hospital discharge was approximated by including only patients who survived 14 days after the index event. We followed patients for recurrent cardiovascular event and the sudden cardiac death (SCD) outcome, with maximum follow-up time set at 8 years as the maximum follow-up time in OSCAR was 7.75 years.

#### Variables and clinical event definitions

Comorbidities were defined by the FHS clinical criteria. Baseline characteristics were taken from the latest clinical examination before the qualifying index event, and past medical history was compiled from all examinations up to the most recent one before the index event.

Cardiovascular events in FHS participants were reviewed and adjudicated by a committee of three physicians. ACS was defined as presence of at least two of the following three criteria: ischemic symptoms, changes in biomarkers reflecting myocardial necrosis, and dynamic ECG changes indicating myocardial ischemia, reflecting the diagnostic criteria of ACS in that period. HF diagnosis was based on the Framingham HF criteria, with presence of at least two major or one major and two minor criteria for HF. SCD was defined as deaths occurring within one hour of symptom onset, with no other probable cause of death other than coronary heart disease^22^. In the FHS cohorts, only fatal SCA cases were included.

## Statistical analysis

We report continuous variables as means (standard deviation) and median (interquartile range) and categorical variables as frequencies and percentages.

Incidence rates are expressed as events per 100 patient-years and calculated by dividing the number of events by the time at risk. For overall incidence rate of SCA, time at risk was taken from the index event to SCA, other mortality or end of follow-up. When calculating the specific incidence rate of SCA in patients with and without a recurrent cardiovascular event, time at risk was defined as follows: for patients without a recurrent event, time at risk extended from the index event to SCA, other mortality, or end of follow-up; for patients with a recurrent event, time at risk started at the time of the first recurrent event and ended with SCA, other mortality, or end of follow-up. The period between the index event and the first recurrence was considered unexposed time and included in the non-exposed group, thereby avoiding misclassification or exclusion of immortal time, which can bias results^23,24^.

We used cumulative incidence functions (CIF) to estimate the cumulative incidence, as traditional Kaplan-Meier curves can overestimate risk in the presence of competing mortality risk^25^.

To assess the risk associated with a recurrent cardiovascular event during follow-up, we used cause-specific Cox proportional hazards models with the recurrent cardiovascular event treated as a time-dependent covariate^23^. In the adjusted Cox model, covariates included were demographics, cardiovascular risk factors, and comorbidities, and a backward stepwise selection procedure was applied, retaining variables with p<0.20. In the OSCAR cohorts, the following variables were tested: age, sex, race and ethnicity, history of smoking, body mass index (BMI), hypercholesterolemia, hypertension, diabetes mellitus, chronic obstructive pulmonary disease (COPD), chronic kidney disease (CKD), peripheral arterial disease (PAD), coronary artery disease (CAD), HF (in the ACS cohort), type of index ACS (STEMI, NSTEMI, UA) and revascularization for index ACS event (in ACS cohort), history of ACS (in HF cohort). In the Framingham cohorts, variables available and tested were age, sex, history of smoking, BMI, hypercholesterolemia, hypertension, diabetes mellitus, HF (in ACS cohort), history of ACS (in HF cohort), and year of index cardiovascular event. For all Cox models, age was treated as a time-dependent variable.

As a sensitivity analysis, we used Fine-Gray sub-distribution hazards models (sHR) to account for competing risks, treating non-SCA mortality as a competing event.

All statistical analyses were performed using SAS version 9.4 (SAS Institute Inc, Cary, NC).

Statistical significance was defined as a two-sided p-value of 0.05 or less.

## Ethical approval

The OSCAR study was approved by the institutional review board (IRB) at CSHS and by the Honest Enterprise Research Broker (HERB) Committee of CSHS’s Enterprise Information Systems (EIS) Research Informatics and Scientific Computing Core (RISCC).

The FHS was approved by the IRB of Boston University Medical Center, and all participants provided written informed consent. The current research was conducted as an ancillary study approved by the FHS Executive Committee.

## Results

### OSCAR discovery cohort

#### ACS cohort - patient population

The ACS discovery cohort comprised 2946 patients, and their baseline clinical characteristics overall and according to outcome can be found in Table 1a. Patients who experienced SCA had a higher prevalence of several comorbidities, including diabetes, CKD and PAD, as compared to those who did not experience SCA. The proportion of patients with a recurrent ACS event was highest among those who subsequently experienced SCA (SCA: 24.6%, other mortality: 14.9%, alive: 11.8%).

**Table 1a).**
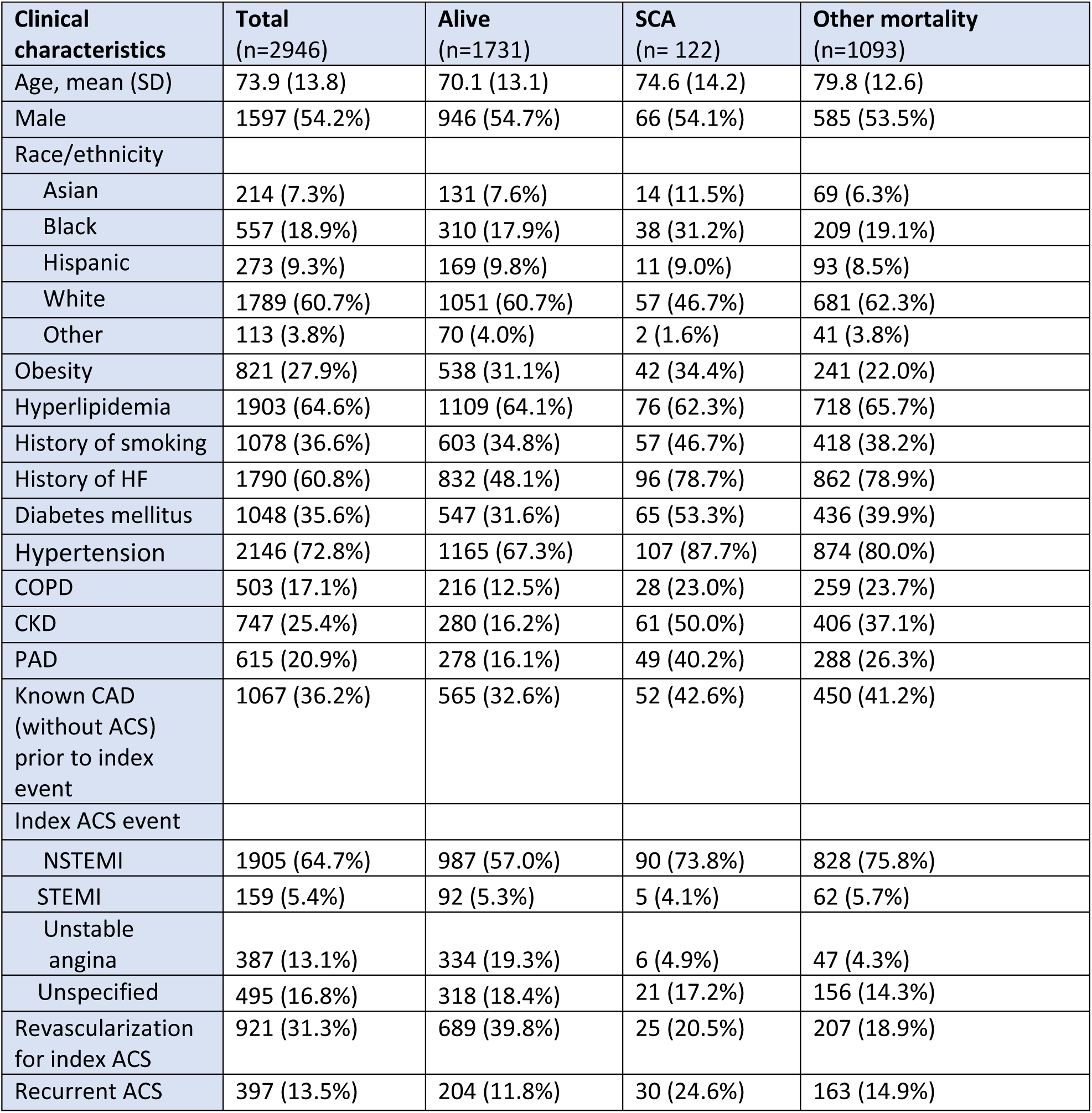
Clinical characteristics according to outcomes in the OSCAR index ACS cohort. Abbreviations: ACS, acute coronary syndrome; CAD, coronary artery disease; CKD, chronic kidney disease; COPD, chronic obstructive pulmonary disease; FHS, Framingham Heart Study; HF, heart failure; NSTEMI, non-ST-segment elevation myocardial infarction; PAD, peripheral arterial disease; SCA, sudden cardiac arrest; STEMI, ST-segment elevation myocardial infarction.

**Table 1b).**
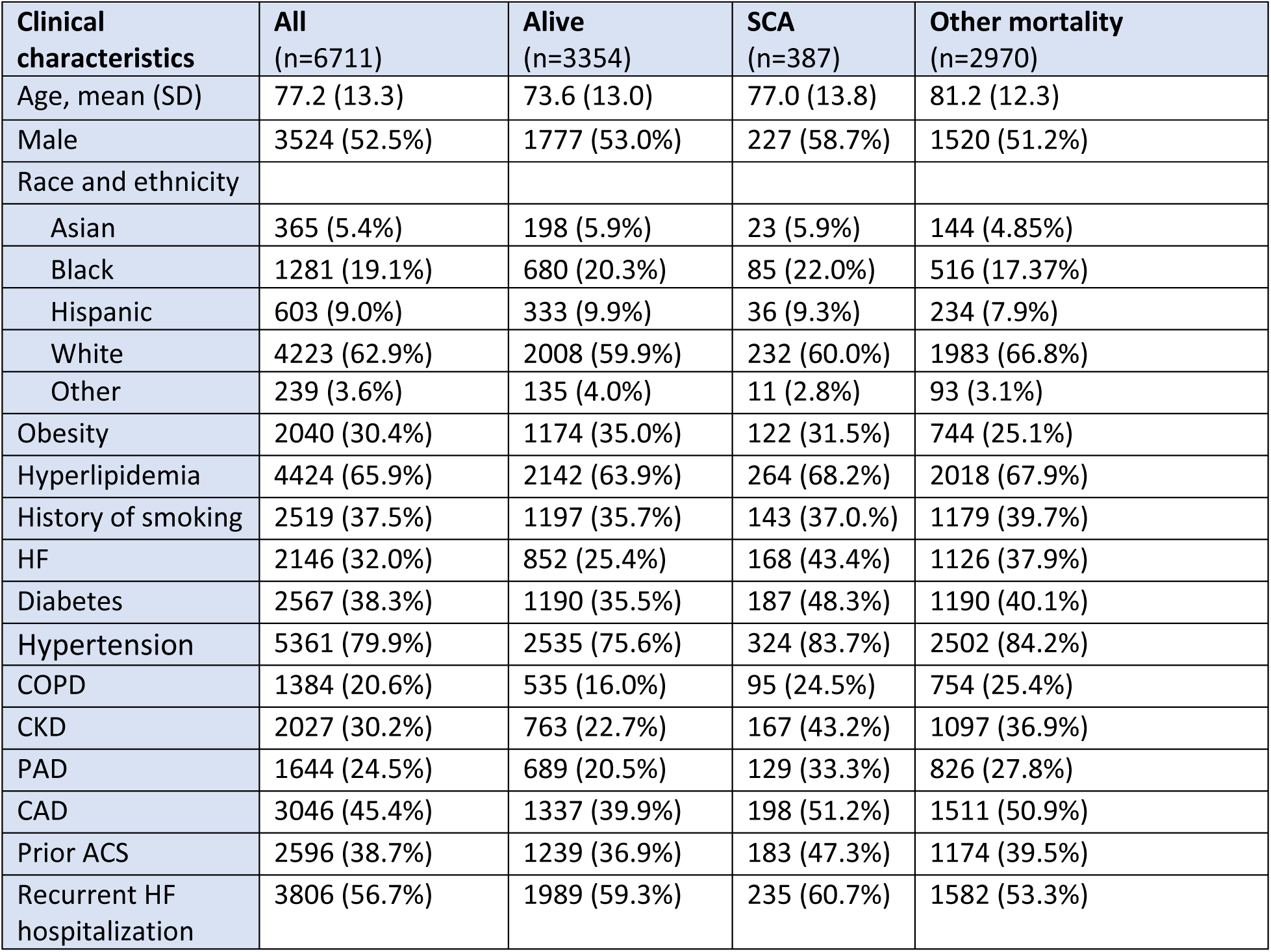
Clinical characteristics according to outcomes in the OSCAR index HF cohort. Abbreviations: ACS, acute coronary syndrome; CAD, coronary artery disease; CKD, chronic kidney disease; COPD, chronic obstructive pulmonary disease; HF, heart failure; PAD, peripheral arterial disease; SCA, sudden cardiac arrest.

#### ACS cohort - Incidence of SCA and influence of recurrent event

Over a median follow-up of 2.3 years, 1215 patients died. In total, 122 patients suffered SCA.

The incidence rate of SCA was 1.53 per 100 patient-years. A recurrent ACS event occurred in 397 (13.5%) patients, and the incidence rate of SCA was greater following a recurrent event compared to no recurrent event (3.70 vs 1.28). The CIF for SCA after a recurrent event and in those without a recurrent event can be seen in Figure 2a. At 5 years of follow-up, the CIF of SCA after a recurrent event was 9.6% and without a recurrent event 3.9%.

**Figure 2a).**
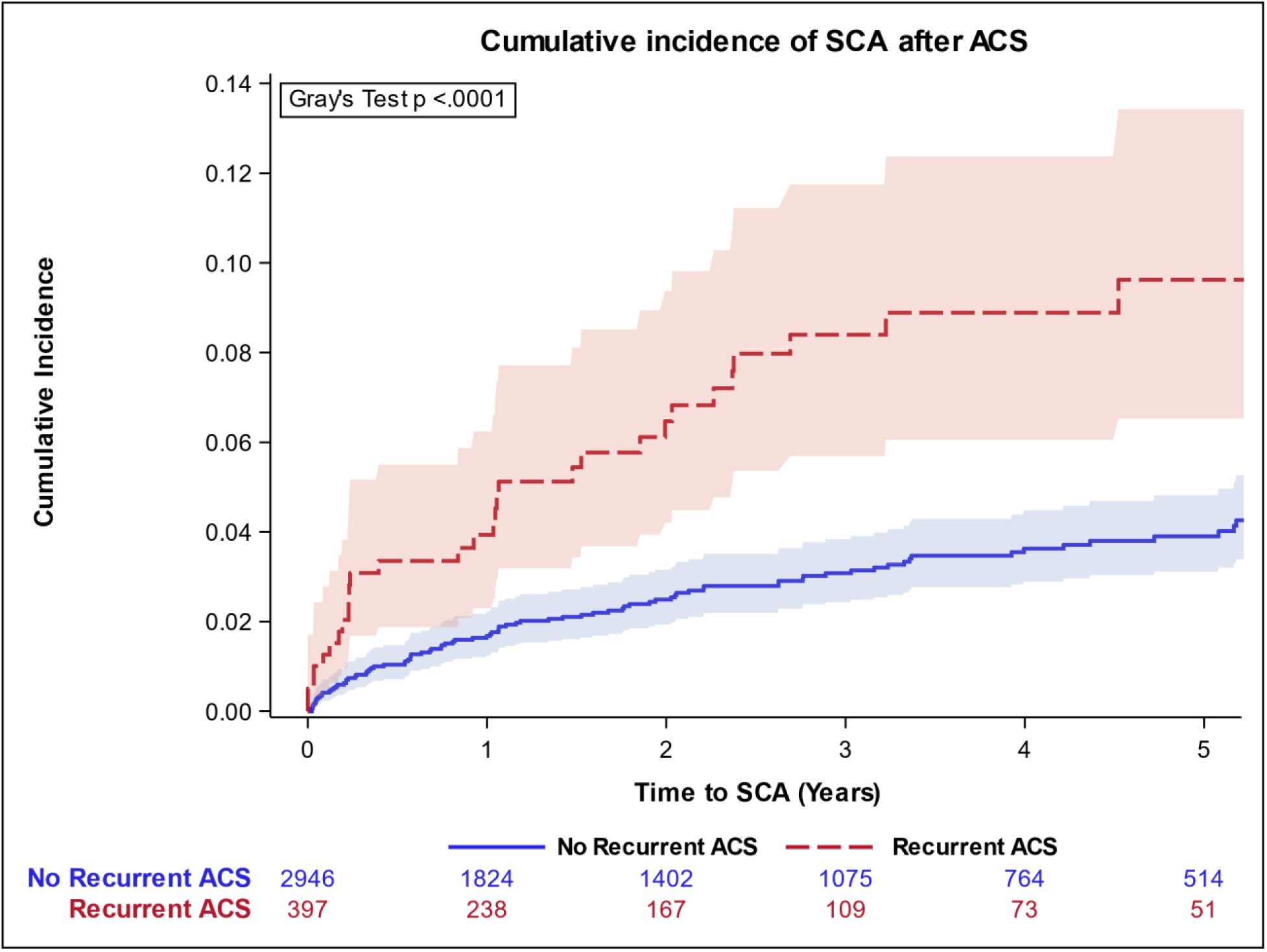
Cumulative incidence function of SCA in patients after a recurrent ACS (red color) and without a recurrent ACS (blue color) in the OSCAR ACS cohort.Abbreviations: ACS, acute coronary syndrome; OSCAR, Observational Study of Cardiac Arrest Risk; SCA, sudden cardiac arrest.

**Figure 2b).**
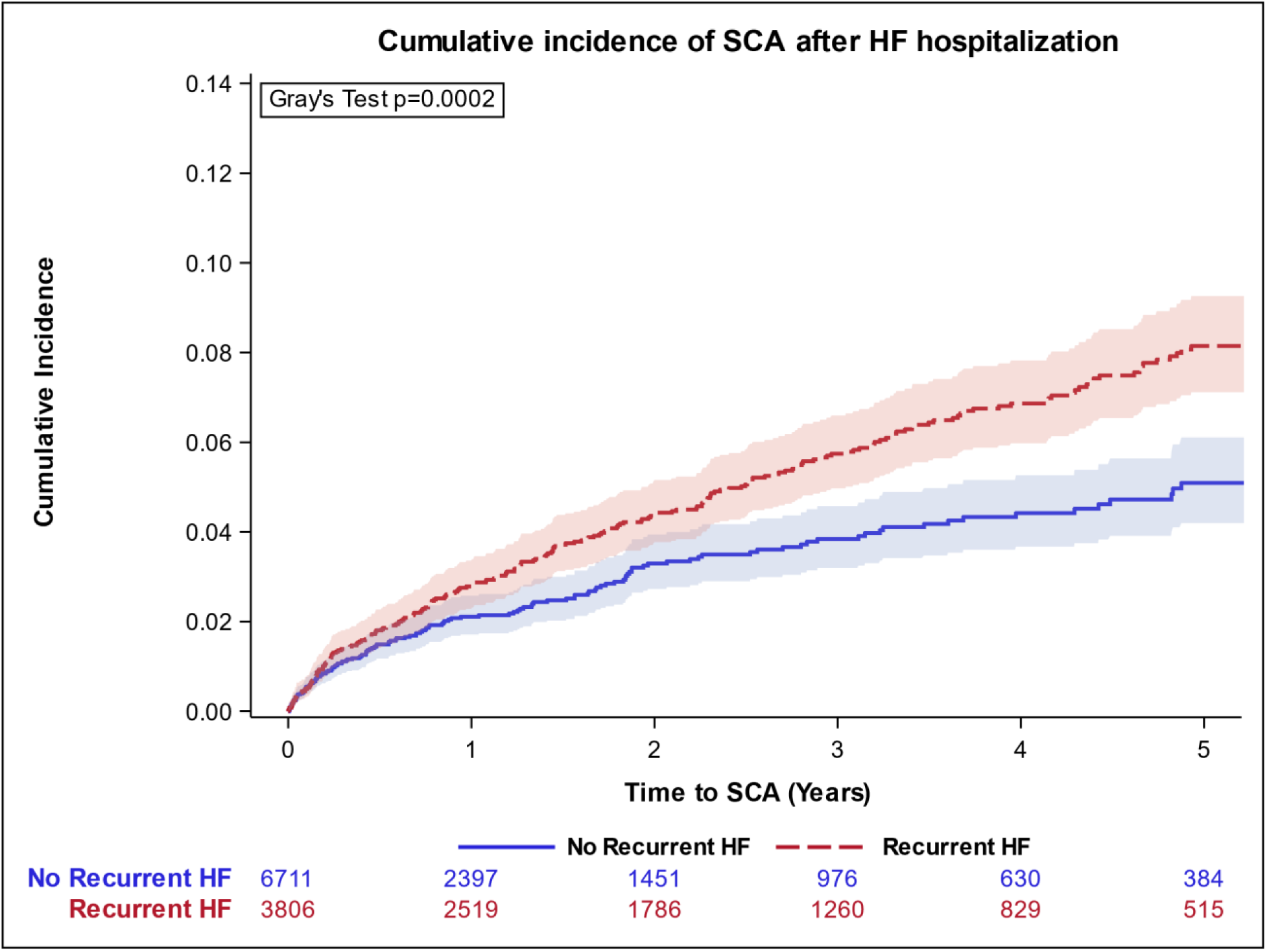
Cumulative incidence function of SCA in patients after a recurrent HF (red color) and without a recurrent HF (blue color) in the OSCAR HF cohort. Abbreviations: HF, heart failure; OSCAR, Observational Study of Cardiac Arrest Risk; SCA, sudden cardiac arrest.

In the cause-specific Cox proportional hazard model with recurrent event as a time-dependent variable, the crude HR for SCA with a recurrent event was 3.51 (2.30-5.36, p<0.0001) (Figure 3). After multivariable adjustment, the HR was 3.15 (2.06-4.83, p<0.0001). In the adjusted model, recurrent event was the variable with the highest associated risk of SCA (Supplementary Table 2).

**Figure 3.**
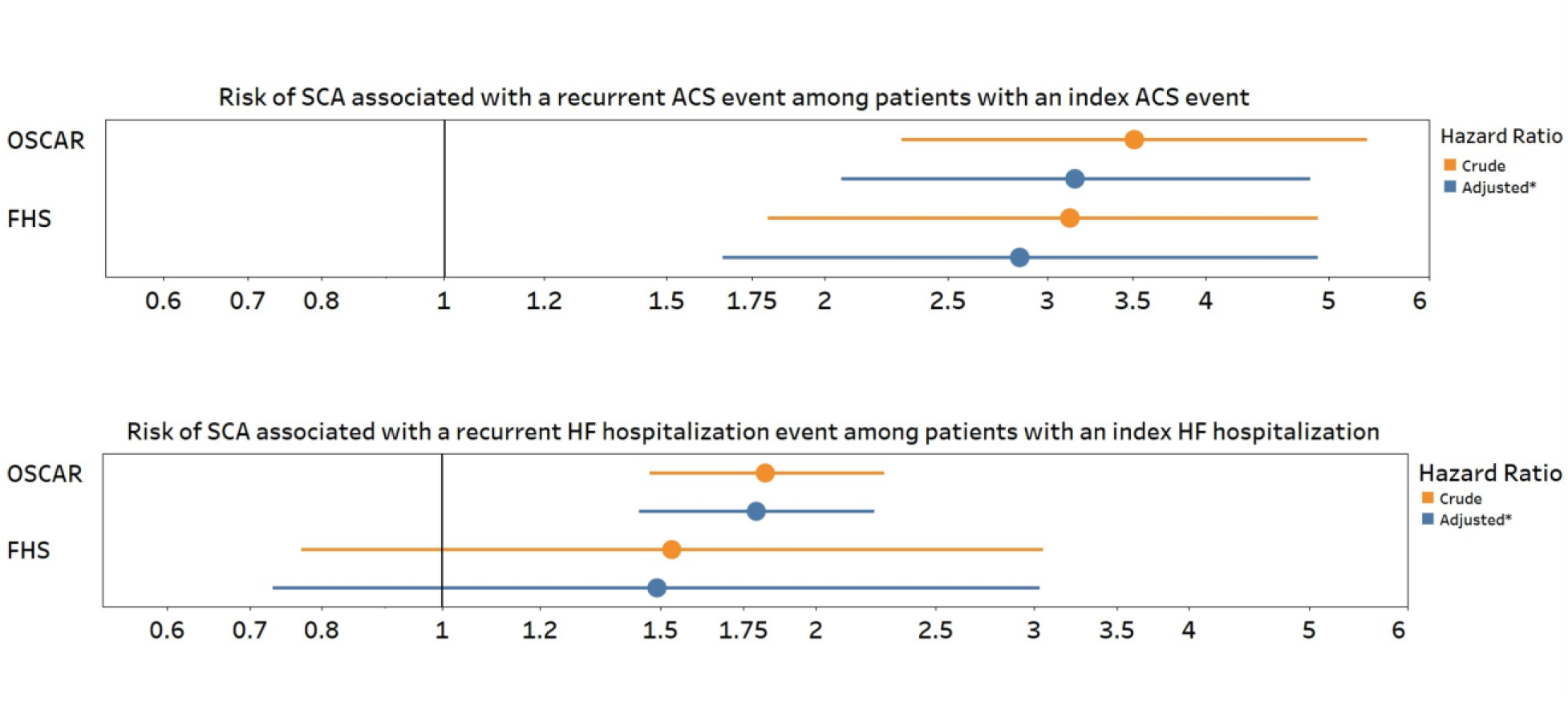
Forest plot of the risk of SCA associated with a recurrent cardiovascular event among patients with an index cardiovascular event. Risks were estimated using cause-specific Cox proportional hazard models, with recurrent cardiovascular events modeled as a time-dependent variable. *Variables included in the adjusted models: OSCAR ACS cohort: age, race and ethnicity, history of smoking, hypercholesterolemia, diabetes mellitus, CKD, PAD, HF, and revascularization for index ACS event; FHS ACS cohort: age, sex, diabetes mellitus, and year of index event; OSCAR HF cohort: age, sex, hypercholesterolemia, diabetes mellitus, CKD, PAD, prevalent HF, and history of ACS; FHS HF cohort: age, sex, and year of index event. Abbreviations: ACS, acute coronary syndrome; CKD, chronic kidney disease; FHS, Framingham Heart Study; HF, heart failure; HT, hypertension; OSCAR, Observational Study of Cardiac Arrest Risk; PAD, peripheral arterial disease; SCA, sudden cardiac arrest.

#### HF cohort - patient population

The HF discovery cohort comprised 6711 patients, whose clinical characteristics both according to outcome and overall can be found in Table 1b. As in the ACS cohort, patients who experienced SCA had higher prevalence of several comorbidities, including diabetes, CKD and ACS compared to those who did not experience SCA. A recurrent HF hospitalization occurred more often in patients who suffered SCA than in those who died of other causes (60.7% vs 53.3%).

#### HF cohort - Incidence of SCA and influence of recurrent event

Over a median follow-up of 2.0 years, 3357 patients died. In total, 387 patients experienced a SCA. The incidence rate of SCA was 2.25 per 100 patient-years. A recurrent HF hospitalization occurred in 3806 (56.7%) patients, and the incidence rate of SCA was higher following a recurrent event compared to no recurrent event (2.63 vs 1.84). The CIF for SCA after a recurrent event and without a recurrent event can be seen in Figure 2b. At 5 years of follow-up, the CIF of SCA after a recurrent event was 8.1% and without a recurrent event 5.1%.

In the cause-specific Cox proportional hazard model, the crude HR for SCA after a recurrent event was 1.82 (1.47-2.27, p<0.0001). After multivariable adjustment, the HR was 1.79 (1.44-2.23, p<0.0001). In the adjusted model, recurrent event had the largest HR among all variables examined (Supplementary Table 2).

### FHS validation cohort

#### ACS cohort

The ACS validation cohort included 1631 patients. Compared to the discovery cohort, the FHS ACS cohort was slightly younger and had a lower comorbidity burden (Table 2a).

**Table 2a).**
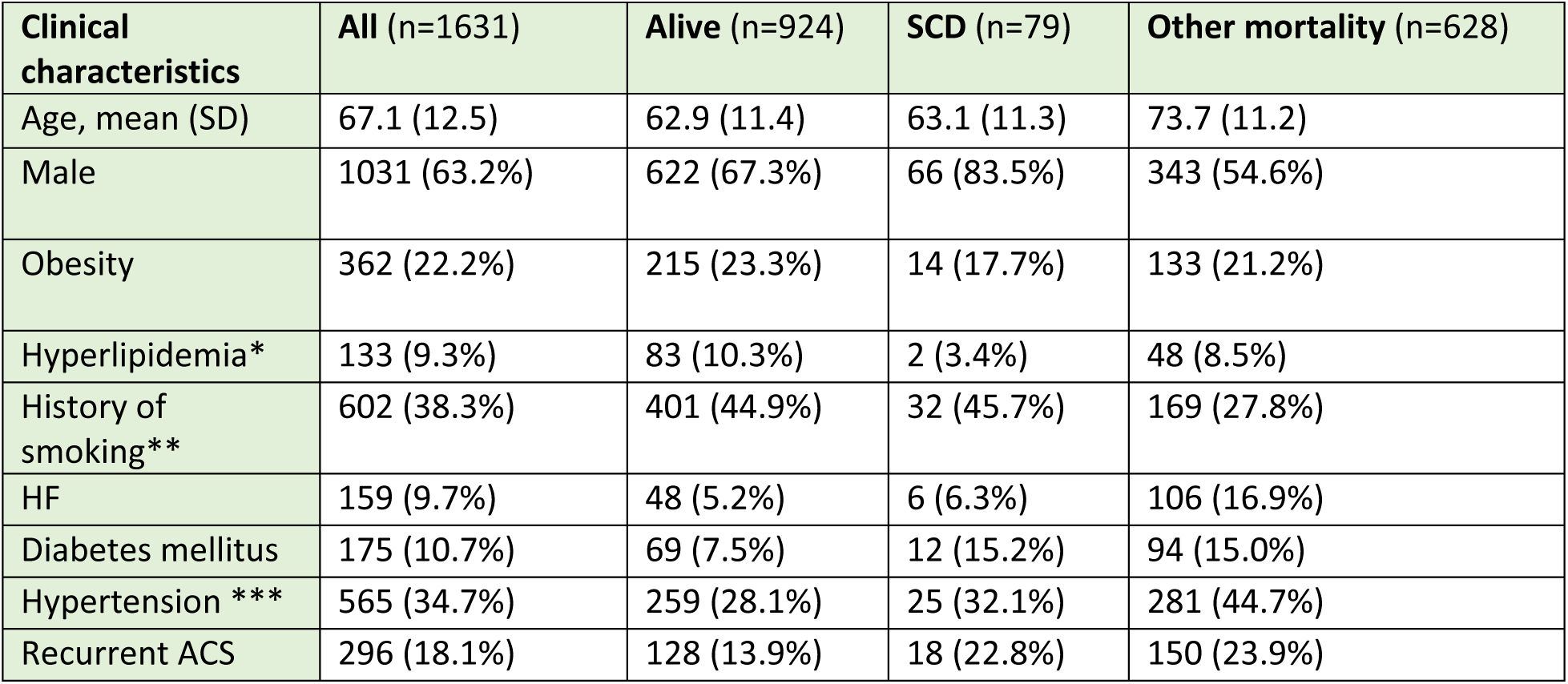
Clinical characteristics according to outcomes in the FHS index ACS cohort. Abbreviations: ACS, acute coronary syndrome; FHS, Framingham Heart Study; HF, heart failure; SCD, sudden cardiac death. * missing n=200. ** missing n=58. *** missing n=3.

**Table 2b).**
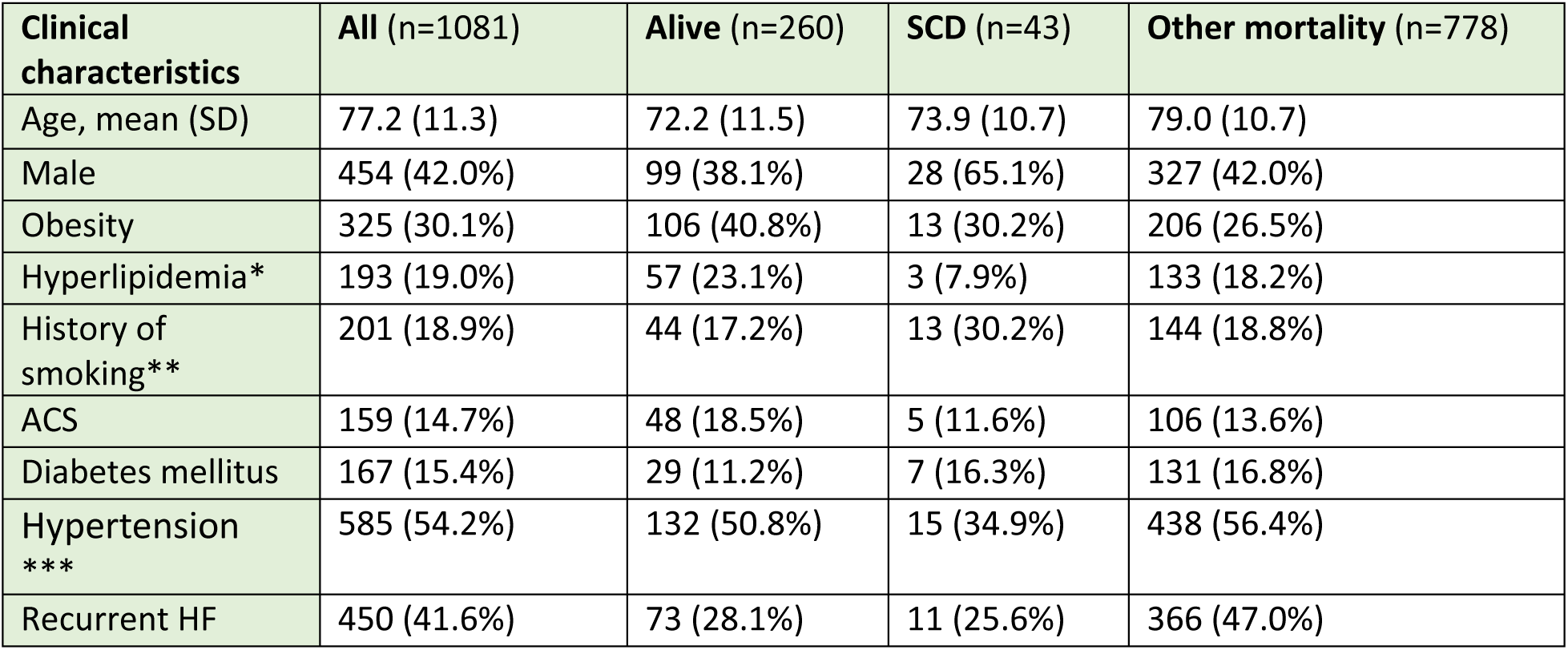
Clinical characteristics according to outcomes in the FHS index HF cohort. Abbreviations: ACS, acute coronary syndrome; FHS, Framingham Heart Study; HF, heart failure; SCD, sudden cardiac death. *missing n=65, **missing n=16, ***missing n=1

Over a median follow-up time of 8.0 years, 707 patients died, of whom 11.2% (n=79) died of SCD. A total of 296 patients experienced a recurrent ACS event. The incidence rate of SCD following a recurrent event was higher than those without a recurrent event (2.09 vs 0.68 per 100 patient-years).

In the cause-specific Cox proportional hazard model, the crude HR for SCD after a recurrent event was 3.12 (95% CI 1.80-5.42, p<0.0001), and the adjusted HR was 2.85 (95% CI 1.66-4.90, p=0.0002).

#### HF cohort

In the HF validation cohort, a total of 1081 patients were included. Baseline characteristics can be found in Table 2b. Overall, the FHS HF cohort had a lower comorbidity burden compared to the OSCAR cohort.

Over a median follow-up time of 3.2 years, 821 patients died, of which 5.2% (n=42) died of SCD. In total 450 (56.7%) patients experienced a recurrent HF hospitalization. The incidence rate of SCD following a recurrent event was higher than without a recurrent event (1.35 vs 0.97 per 100 patient-years).

In the cause-specific Cox proportional hazard model, the crude HR for SCA after a recurrent HF hospitalization was 1.53 (95% CI 0.77-3.05, p=0.236), and the adjusted HR was 1.49 (95% CI 0.73-3.03, p=0.273).

### Sensitivity analysis

A competing-risks sensitivity analysis was conducted in the OSCAR cohorts, in which we calculated the sHR for SCA treating other-cause mortality as a competing risk event. In the ACS cohort, the sHR for SCA associated with a recurrent event was 3.13 (95% CI 2.03-4.83, p<0.0001), and in the HF cohort 2.02 (95% CI 1.64-2.50, p<0.0001).

## Discussion

In this observational prospective cohort study of patients with a first-time ACS or HF hospitalization, recurrent cardiovascular events were associated with significantly higher risk of SCA. Specifically, in the index ACS cohort, a recurrent ACS event was associated with more than a threefold higher risk of SCA. Similarly, in the index HF cohort, a recurrent HF hospitalization was associated with a more than 80% higher risk of SCA. These findings were successfully validated with the same level of increased risk in the external validation cohorts from the Framingham Heart Study, although not reaching significance level for the HF cohort. To our knowledge, this is the first study to examine recurrent ACS or HF events as a potential dynamic risk marker for SCA in a contemporary, real-world population.

Predicting SCA remains a significant challenge. Both ACS and HF are well-established major risk factors for SCA, as they can lead to cardiac electrical and structural remodeling that increases susceptibility to malignant arrythmias. It is also recognized that SCA risk evolves over time, influenced by changes in the underlying substrate as well as transient triggers^7^. Among survivors of a first cardiovascular event, the risk of recurrent events, including new ACS episodes or HF hospitalizations, as well as cardiovascular and all-cause mortality, is elevated^9–11^.

Moreover, mortality risk is further increased among those who do experience a recurrent event^12^. However, the specific risk of SCA in this context has not been well characterized. In line with our results, a few post hoc analyses of RCTs have indicated a higher risk of SCA following recurrent ischemic events^13–15^, whereas the risk after recurrent HF hospitalizations has not been previously reported.

### Recurrent ACS and risk of SCA

In the OSCAR cohort of patients who experienced a first ACS, 13.5% went on to experience a new ACS event over a median follow-up period of 2.3 years. This rate of recurrent events aligns with findings from another large, US-based EHR medical claims cohort of a contemporary ACS population^11^.

In both the ACS cohorts from OSCAR and FHS, the incidence rate of SCA was higher following a recurrent ACS event compared to those without a recurrent event. An EHR-based study conducted in Finland on ACS patients undergoing invasive evaluation similarly reported a higher cumulative incidence of SCA among patients who experienced a recurrent MI as compared to those who did not (10.4% vs 6.2% at 12 years follow-up)^15^. Furthermore, in a post-hoc analysis of the IMPROVE-IT trial, a RCT involving stabilized ACS patients, the risk of sudden death was likewise higher after a recurrent cardiovascular event (composite of ACS, stroke, and hospitalization for HF) than in those without such an event (1.45 vs 0.27 per 100 patient-years)^14^.

In the current study, survival analyses showed that a recurrent ACS event during follow-up was associated with a threefold higher risk of SCA in both the discovery and validation cohort. A sensitivity analysis, accounting for competing mortality risk, supported these findings with similar results. These observations are consistent with results from the IMPROVE-IT study, as well as in a pooled RCT analysis of NSTEMI patients, reporting a three times higher risk of sudden deaths in patients that experienced a recurrent cardiovascular event^13^. Further, in a post-hoc analysis of the VALIANT RCT involving patients with MI and HF, landmark analyses for different time intervals showed that recurrent MI, HF hospitalization, or hospitalization for any cause were consistently associated with a significantly increased risk of SCD^26^. Unlike these earlier published studies, the current study included all patients with a first-time ACS event, specifically examined recurrent ACS events as the exposure (rather than a composite exposure variable), and used comprehensively adjudicated SCA outcomes. This enhances the clinical relevance and applicability of our findings. A prior study ^27^ reported that recurrent ischemic events were not significantly associated with increased coronary artery related SCD risk (HR 1.26, p=0.09). However, the Adabag study was not contemporaneous (1979 to 2005) and was limited to Olmsted County, MN. The current study was conducted in a multi-ethnic population under modern treatment patterns, making the findings more generalizable and relevant for today’s clinical practice. The competing risk analysis performed in the current study further strengthens the robustness of the findings.

### Recurrent HF and risk of SCA

In the OSCAR cohort of patients experiencing a first HF hospitalization, 56.7% of patients experienced a recurrent HF hospitalization over a median follow-up time of 2.0 years. Reported proportions in the literature vary depending on HF severity, phenotype, and comorbidity burden, but consistently indicate that recurrent hospitalizations are common^28,29^.

Several studies have reported a significantly increased risk of overall mortality in patients with repeated HF hospitalizations^28,30–33^. To our knowledge, no prior studies have specifically examined difference in SCA risk between patients who do or do not experience a recurrent HF hospitalization. Among patients with HF, some studies have suggested a higher risk of malignant arrhythmias in those with recurrent HF hospitalizations. A post hoc analysis of the ICD arm of MADIT-ll, including patients with MI and HF with reduced ejection fraction (HFrEF), reported that HF hospitalization was associated with an HR of 2.5 for appropriate ICD shocks. Similarly, a pooled analysis of RCTs involving patients with both ischemic and non-ischemic cardiomyopathy and HFrEF reported that HF hospitalization was associated with an HR of 1.79 for ventricular tachyarrhythmias. Although these studies were restricted to patients with HFrEF and focused on ventricular tachycardias as outcomes, they provide a similar signal to our current study. In the current study, recurrent HF hospitalizations were associated with an 82% increased risk of SCA, and this association remained consistent in a sensitivity analysis that accounted for other causes of mortality as a competing risk. Although the associated increased risk was comparable in OSCAR and FHS, the finding was non-significant in the FHS, likely due to the smaller sample size and fewer SCD events.

### Dynamic risk trajectories

Across both the index ACS and HF cohorts, the elevated risk of SCA following a recurrent cardiovascular event was consistent across all Cox models (crude and adjusted) and in competing-risk analyses, underscoring the robustness of the association. These findings support a dynamic-risk framework, wherein temporal changes in clinical characteristics during follow-up provide incremental prognostic information beyond baseline factors^7^. The pattern aligns with emerging evidence that longitudinal trajectories, rather than a single time-point measurement or static risk assessment, offer superior risk stratification. We have previously reported that ECG remodeling over time is itself a significant risk marker for SCA, reinforcing the value of dynamic assessment^6^. Future investigation should extend this approach by identifying additional dynamic risk markers and potentially developing risk-prediction models that incorporate these factors to improve SCA prediction.

### Strengths and Limitations

The OSCAR study enables ascertainment of a large, contemporary, real-world cohort of patients with index ACS or index HF hospitalizations, and the unique integration of both EHR and EMS data facilitates a robust and comprehensive adjudication process for SCA. Importantly, the main findings from OSCAR were replicated in the well-established FHS, strengthening the validity and generalizability of the results.

While both studies support the main findings, there are notable differences between the OSCAR and FHS cohorts. OSCAR is an EHR-based cohort of patients that receive regular care within an advanced healthcare system, while FHS in contrast is a community-based cohort designed to identify cardiovascular risk factors in included participants who were unselected for conditions at baseline. In addition, the FSH was initiated in an earlier era with different clinical practices and treatment options for ACS and HF. This could potentially explain why the OSCAR cohorts were older, had higher comorbidity burden and different absolute event rates.

Additionally, OSCAR SCA cases included survivors (5.1% of total SCA cases), while FHS included only fatal SCD. Despite these differences, the consistency of the primary finding of higher risk of SCA after a recurrent cardiovascular event across cohorts reinforces the robustness of the findings.

In the OSCAR study, recurrent cardiovascular events may have been missed if they occurred at institutions outside of Cedars-Sinai Health System, and cases of SCA could have been missed if they occurred outside LA County. In the FHS, the cohorts are composed primarily of individuals of European ancestry residing in New England, which may limit generalizability of the findings to other racial, ethnic and geographic populations. For both OSCAR and FHS, the possibility of residual confounding cannot be excluded, and as both studies are observational in nature, associations can be described but causality cannot be established.

There is a potential risk of immortal time bias as the exposure of interest occurs during follow-up. To mitigate this risk, we employed survival analyses with time-dependent covariates, ensuring that person-time was appropriately classified according to exposure status.

## Conclusion

In a contemporary observational, prospective health system cohort study of patients with a first-time ACS or HF hospitalization, recurrent cardiovascular events were associated with a significantly increased risk of SCA. These findings were externally validated in the community-based Framingham Heart Study. These findings highlight the importance of incorporating recurrent cardiovascular events into dynamic risk assessment frameworks to inform targeted management and prevention strategies for SCA in diverse clinical settings.

## Data Availability

De-identified data may be made available from the corresponding author upon reasonable request and with permission from Cedars-Sinai Medical Center and the Framingham Heart Study.

## Acknowledgments

The authors would like to thank the Los Angeles County EMS for their significant contributions, and all participants and members of the FHS.

## Non-standard Abbreviations and Acronyms

ACS: Acute coronary syndrome
CAD: Coronary artery disease
CKD: Chronic kidney disease
COPD: Chronic obstructive pulmonary disease
EHR: Electronic health record
FHS: Framingham Heart Study
HF: Heart failure
ICD: Implantable cardioverter-defibrillator
LVEF: Left ventricular ejection fraction
NSTEMI: Non-ST-segment elevation myocardial infarction
MI: Myocardial infarction
OSCAR: Observational Study of Cardiac Arrest Risk
PAD: Peripheral arterial disease
RCT: Randomized controlled trial
SCA: Sudden cardiac arrest
SCD: Sudden cardiac death
STEMI: ST-segment elevation myocardial infarction

